# Information nudges for influenza vaccination: Evidence from a large-scale cluster-randomized controlled trial

**DOI:** 10.1101/2021.04.27.21255975

**Authors:** Lauri Sääksvuori, Cornelia Betsch, Hanna Nohynek, Heini Salo, Jonas Sivelä, Robert Böhm

**Author notes:** Address correspondence to: Lauri Sääksvuori.

## Abstract

**Background:** Vaccination is the most effective means to prevent the spread of infectious diseases. Despite the proven benefits of vaccination, complacency, constraints, and lacking confidence keep many people away from getting vaccinated. This study investigates how written reminders with varying information contents to address vaccine hesitancy affect influenza vaccination coverage in two large and representative samples of older adults.

**Methods:** We implemented a large-scale cluster-randomized controlled trial in Finland. The study included the entire elderly population (≥ 65 years of age) in two culturally and geographically distinct regions with a historically low (31·8%, N = 7398) and high (57·7%, N = 40727) influenza vaccination coverage. Participants were randomized before the influenza season 2018 – 2019 into three treatments: (i) no information letter, (ii) a standard information letter, reminding recipients about the individual benefits of vaccination, and (iii) a tailored information letter, reminding recipients about the additional social benefits of vaccination due to herd effect. The impact of varying information treatments on influenza vaccination coverage was measured using individual-level administrative health records.

**Findings:** Our results showed that a low-cost and scalable information intervention relying on individually mailed letters increased influenza vaccination coverage by 6·4 percentage points (95% CI: 4·1 to 8·8). The effect was particularly large among individuals with no prior influenza vaccination (8·8 pp, 95% CI: 6·5 to 11·1). Moreover, we observed a substantial positive effect (5·3 pp, 95% CI: 2·8 to 7·8) among the most consistently non-vaccinated individuals who had not received any type of vaccine during the previous nine years. There were no cross-vaccine spillovers to other age-appropriate vaccines. Our results further suggest that there was no difference in influenza vaccination coverage between the standard letter and the tailored letter that informed individuals about the social benefits of vaccination (0·2 pp, 95% CI: - 0·1 to 1·3).

**Interpretation:** Sending information letters is an effective and easily scalable low-cost intervention strategy to increase vaccine uptake in an elderly population. Communicating the social benefits of vaccination in addition to individual benefits does not enhance influenza vaccine uptake. The effectiveness of behavioral interventions aiming to improve vaccination coverage crucially depends on the prior vaccination history of the target population. These findings have meaningful implications for public health authorities who implement vaccine communication strategies to enhance vaccine uptake and aim to curb the spread of infectious diseases.

**Funding:** The authors received no external funding for this work. The costs of preparing (e.g. printing the letters and acquiring envelopes) and mailing the letters (postal fees) were paid by the Finnish Institute for Health and Welfare and the City of Espoo.

## Introduction

Vaccination has historically made an enormous contribution to global health. Large-scale vaccination programs continue to reduce morbidity and mortality due to numerous infectious diseases and create the backbone of health security strategies across the globe. However, increasing levels of vaccine hesitancy – delay in acceptance or refusal of vaccines despite the availability of vaccination services – threatens to reverse the progress made in halting vaccine-preventable diseases.^1-3^ In 2019, the World Health Organization (WHO) declared vaccine hesitancy as one of ten biggest threats to global health. Understanding how to improve vaccine uptake and overcome the different behavioural mechanisms of vaccine hesitancy is important, not only to improve the current vaccination coverage but also to secure high coverage of any new vaccine.

Several behavioral factors have been identified as relevant predictors of non-vaccination. These factors include confidence (lack of trust in safety and effectiveness of vaccination), complacency (lacking risk perception), constraints (perceived or actual structural and behavioral barriers such as forgetting or difficulties in access), calculation (engagement in extensive information searching), and lacking collective responsibility (willingness to protect others).^4,5^ Despite the accumulating evidence about the psychological antecedents of vaccination decisions and the development of validated measures to understand vaccine hesitancy, there is little causal evidence about the behavioral mechanisms that drive vaccine hesitancy. Moreover, there is little causal evidence about the effectiveness of scalable low-cost behavioral interventions that can be used to increase vaccine uptake among large population groups.

Beside the importance for public health, vaccination is a canonical example of positive behavioural externalities.^6,7^ Vaccinations do not only incur individual benefits through direct protective effects, but they affect the community at large through indirect effects that reduce the risk of spreading the disease to others and build up herd effect.^8^ These externalities provide simultaneously motivation for pro-social vaccination to protect unvaccinated individuals and incentives to free-ride on the vaccination of others to avoid the costs and potential risks of vaccination. Existing empirical research has shown that educating individuals about the social benefits of vaccination can increase their intentions to vaccinate.^9,10^ Consequently, highlighting the social benefits of vaccination due to herd effect appears to be a promising candidate to promote vaccination uptake in large population groups.

This study aims to understand the behavioral determinants of individual vaccination decisions and improve the uptake of influenza vaccination. The focus is on influenza vaccination among the elderly population where the gap between the vaccination target and vaccination coverage is particularly large.^11^ However, the problem of low vaccination coverage is not limited to seasonal influenza vaccines but observed across a wide range of vaccines, inflicting a widespread public health concern and substantial economic costs.^12^

This study tests the effect of written reminders – distributed via regular mail – on influenza vaccination coverage using a large-scale cluster-randomized controlled trial. Comprehensive nationwide health care records on influenza vaccination coverage following our intervention period serve as the main outcome variable and determine the effectiveness of information treatments.

The use of administrative health care records as the main outcome variable has several key advantages. First, we are not restricted to study vaccination intentions or self-reported vaccination outcomes, but can objectively measure whether and when the vaccination occurred. Second, individuals residing in the study regions were not aware that different letters were sent to eligible individuals. The written letters themselves did not make any reference to experimental variation in wording. As a result, the generalizability of our results is not limited by the common concern that experimental results based on voluntary participation do not generalize to the target population of interest that includes the population that was not aware of the experiment or did not volunteer for the experiment when offered the opportunity. Third, the use of administrative patient records as the main outcome variable enabled a sample size that is an order of magnitude larger than in typical randomized controlled trials that require the use of survey instruments to measure outcome variables. Finally, administrative patient records on all vaccinations enabled us to measure potential behavioral spillovers to other age-appropriate vaccinations.

## Methods

### Study design

We implemented a cluster-randomized controlled trial in two geographically and culturally distinct communities in Finland. The trial had two active treatment arms. The first treatment, Individual (I), highlighted the individual benefits of vaccination. The second treatment, Individual and Social (I + S), highlighted the social benefits of vaccination in addition to the individual benefits. There was a control group without any intervention in the Western region. There was no control group without any intervention in the Southern region. The local health care authority had previously sent annual reminder letters to the entire population aged 65 and above residing in the Southern region. It was considered to be a good and ethical research practice that the intervention would not leave anyone without the information that they would have received in the absence of the intervention. The study was implemented in a partnership with the local health care authorities in both regions.

The letters varied the information content of individual invitation letters (original letters are available in Appendix A). The individual benefit letter contained basic information about the severity of influenza symptoms, seasonal influenza vaccination, the availability of vaccination (locations and dates to receive the vaccination) and instructions about how to book an appointment for the vaccine administration. The experimental variation between the different letters was related to the description of the social benefit of the vaccination. In addition to the individual benefit letter, the individual and social benefit letter contained information about herd effect that read as follows:

“*Your decision to vaccinate does not only protect you but others as well. Your vaccination may protect small children whose immune system is still developing. You will be able to protect your loved ones who will not be able to vaccinate themselves. Your vaccination may prevent the spread of influenza viruses. Thus, the whole society benefits from your decision to vaccinate*.*”*

The analysis sample includes only individuals living either in single- or two-person households. The original sample included housing units with more than two-persons aged 65 and above (e.g. nursing homes). Yet, we excluded housing units with more than two-persons aged 65 and above from the analysis sample because these were often private nursing homes that provide seasonal influenza vaccination to all residents as part of their care plan. There were no other scientific, ethical, or economic reasons to exclude any individuals who met the specified inclusion criteria. Moreover, since the marginal costs of including additional individuals in these types of information interventions is very low, it is worthwhile to maximize the statistical power to detect even potentially small effect sizes. Notably, low-cost interventions with small effect sizes can be highly cost-effective and welfare enhancing investments.

### Study population

The study took place in two regions with widely varying baseline vaccination coverage to test the effect of different letters in two different contexts and obtain information about the potential generalizability of the findings across populations with differing baseline vaccination coverages and socio-economic characteristics. We focused on elderly adults as they belong to a risk group with higher morbidity and mortality from influenza viruses than the prime working age population.^13,14^

The study population included all individuals who were born in the year 1953 or before and resided in the two target regions on June 01, 2018. These two regions represent populations with different historical influenza vaccination coverage (Figure 1). The Western region on the West coast of Finland is a rural region that contains five independent municipalities (Maalahti, Korsnäs, Närpiö, Kaskinen and Kristiinankaupunki). The region has a single public provider of primary health care services that is co-owned by the municipalities. This region has low influenza vaccination coverage among the 65 years and older age group (31·8% during the influenza season 2017 – 2018) compared to the national average (47·7% during the influenza season 2017 – 2018). The Southern region covers the city of Espoo which is the second largest city in Finland. The population in the Southern region belongs to the inner urban core of the Helsinki metropolitan area and has one the highest rates of influenza vaccination coverage among the 65 years and older age group in Finland (57·7% during the influenza season 2017 – 2018).

**Figure 1:**
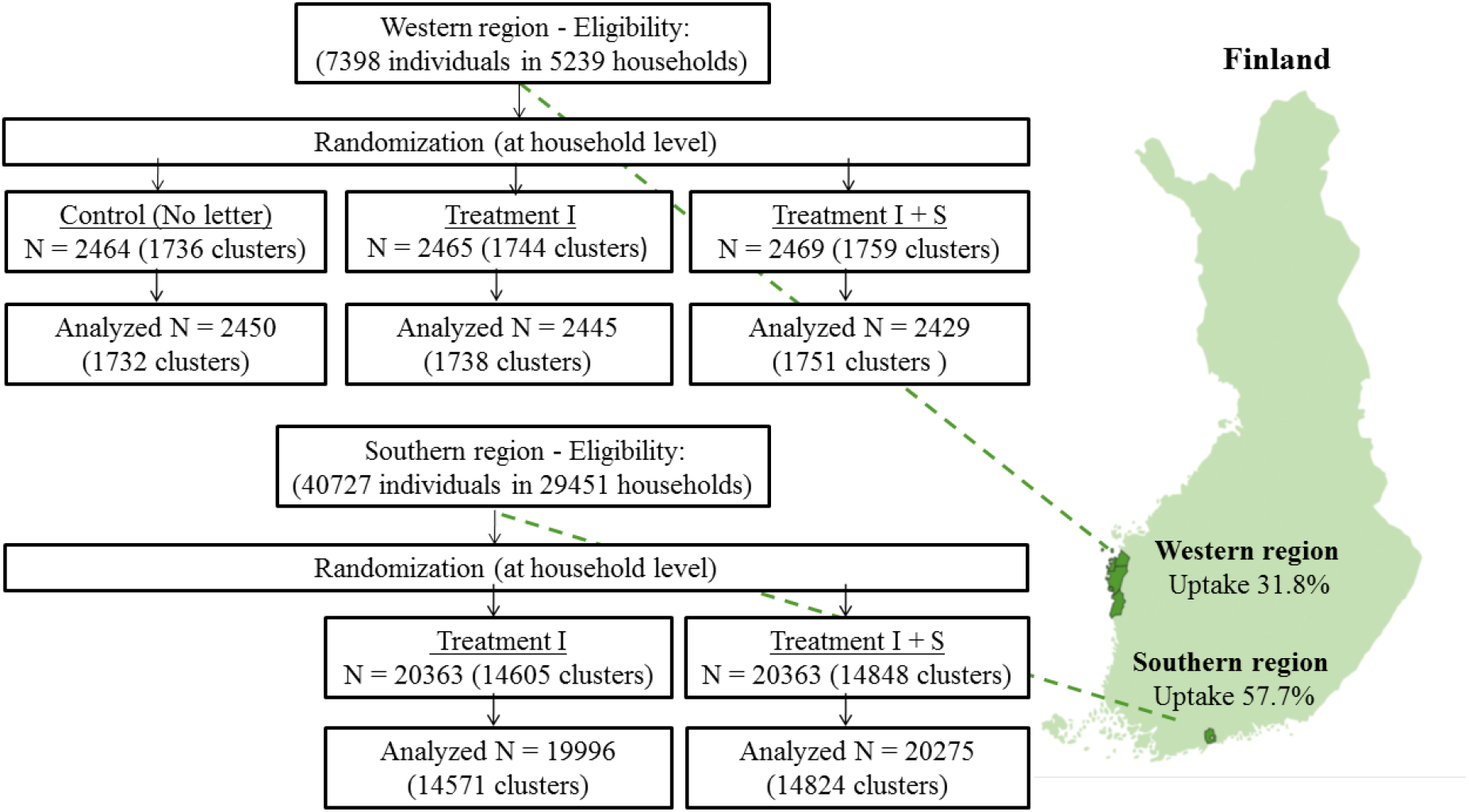
Study regions and randomization scheme

### Randomization and masking

We used the Finnish Population Register to identify the name and postal address of individuals who fulfilled the eligibility criteria (age and place of residence). Randomization took place at the household (cluster) level to avoid sending letters with different contents to the same household members (Figure 1). The randomization at household level was implemented using unique apartment IDs and was performed using a computer-generated randomization code written by the authors (Appendix B). The universe of individuals who were born in 1953 or before and resided in the Western region on June 01, 2018 (N = 7398) was randomized to three equally large treatment arms: (i) No letter, (ii) Individual, and (iii) Individual and Social. The universe of individuals who were born in year 1953 or before and resided in in Region B on June 01, 2018 (N = 40727) was randomized to two equally large treatment arms: (i) Individual, and (ii) Individual and Social.

Individuals residing in the study regions and belonging to the target group were not aware that they were studied. The written letters themselves did not make any reference to experimental variation in wording. The nurses administering the influenza vaccination during the follow-up period were not aware that different letters were sent to eligible individuals. We had neither any direct contact to the recipients of the mailed letters nor to the nurses administering the influenza vaccinations in the target regions. The study protocol was approved by the Finnish Institute for Health and Welfare Institutional Review Board (Decision Number: THL/1444/6.02.01/2018).

### Outcomes

The impact of different letters on vaccination coverage was measured at individual-level using administrative health records. The Finnish National Vaccination register contains nationwide records of all vaccinations given at public health care units in Finland since 2009.^15^ We used data on the lot number of the vaccine to identify the vaccine type and a time stamp to identify the date of vaccine administration. The main outcome variable was having received an influenza vaccination during a four-month follow-up period (from October 18, 2018 to March 18, 2019), coded as one, otherwise coded as zero. Moreover, we used data on prior vaccination history to study the potential heterogeneity of average treatment effects.

### Procedure

We used the Finnish Population Register to identify the name and postal address of individuals who fulfilled our eligibility criteria (age and place of residence). Importantly, an extract from the Population Register contains an individual identifier (social security number) that can be used to match the received letter with complete vaccination records from the Finnish National Vaccination Register. All letters were sent via regular post to the eligible individuals on October 17, 2018. All letters were double sided and written both in Finnish and Swedish to take into account bilingual study populations. The final dataset was produced using individual identifiers (encrypted social security numbers) that enabled us to merge the population register data to the administrative vaccination records. The final dataset analysed by the authors did not contain any information that would allow direct identification of personal information.

### Trial registration

As this study spans multiple disciplines, we pre-registered the experimental design and submitted the pre-analysis plan to multiple registries: the U.S National Library of Medicine Registry for clinical trials (clinicaltrial.gov, trial number: 240317), the American Economic Association Registry for randomized controlled trials (trial number: AEARCTR-0003520), and aspredicted.org (trial number: #15682).

### Statistical analysis

Taking into account the likely correlation of outcomes within (two-person) households, randomization at household level and prior baseline vaccination rate of roughly 32% in the Western region, we computed that a sample size of 2465 individuals per treatment, divided into 1750 clusters with an assumed intracluster correlation of 0.5, was sufficient to obtain 80% power for a 5% (two-sided) level test for at least 3·5 percentage point difference in the probability of receiving an influenza vaccination between any two treatments. Combining active treatment arms to estimate the impact of an information letter per se allows detecting even smaller effects with 80% power.

The study population in the Southern region was divided into two equally large treatment groups. Following the same assumptions as above and taking into account the prior baseline vaccination rate of 58% in the Southern Region, we computed that a sample size of 40271 individuals, divided into two treatments and 29395 clusters, was sufficient to obtain 80% power for a 5% (two-sided) level test for at least 1·5 percentage point difference in the probability of receiving an influenza vaccination between the two treatments. More comprehensive power calculations that vary in the statistical power and assumed intracluster correlation are available in the online Appendix C.

To assess the impact of an information letter per se, we combined the individual (I) and the individual and social (I + S) letter and estimated the effect of receiving any letter on influenza vaccination coverage. We estimated statistical models using linear probability estimation where the coefficients represent marginal effects. We used linear probability models for simplicity and easy interpretation of coefficient values. Appendix D provides results using logit models and multilevel mixed effect linear models with an error structure that allows for cluster level heterogeneity (random effect) at the household level. These alternative regressions models provide extremely similar results. As pre-registered, the regressions did not include any control variables.

To examine the impact of varying types of information letters, we separately estimated the effects of the individual (I) letter and the individual and social (I + S) letter on influenza vaccination coverage. We estimated also these statistical models using linear probability estimation where the coefficients represent marginal effects. In all regression models, we used standard errors that are clustered at the household level.

## Results

### Population and baseline characteristics

Our sample included all citizens aged 65 and above living in single- or two-person households in the two target regions. Table 1 displays baseline characteristics across the regions and treatments, showing large differences in the proportion of previously vaccinated individuals between the Western and Southern regions. The coverage of influenza vaccination among our target population was 31·8% in the Western region and 57·7% in the Southern region at the end of the influenza season 2017 – 2018. Notably, the differences in vaccination coverage were not limited to influenza vaccination. The proportion of individuals having received any vaccination during the previous nine years was 72·5% in the Western region and 81·0% in the Southern region.

**Table 1:** Descriptive statistics by study region and treatment (analysis sample)

|  | Western region |  |  | Southern region |  |
| --- | --- | --- | --- | --- | --- |
|  | No letter<br>(N = 2450) | Treatment I<br>(N = 2445) | Treatment I+S<br>(N = 2429) | Treatment I<br>(N = 19996) | Treatment I+S<br>(N = 20275) |
| Influenza vaccination –<br>previous season | 787<br>[32.1%] | 818<br>[33.5%] | 724<br>[29.8%] | 11567<br>[57.8%] | 11683<br>[57.6%] |
| Influenza vaccination –<br>any year | 1097<br>[44.8%] | 1113<br>[45.5%] | 1033<br>[42.4%] | 14280<br>[71.4%] | 14292<br>[70.5%] |
| Any vaccination | 1752<br>[71.5%] | 1809<br>[74.0%] | 1747<br>[71.9%] | 16243<br>[81.2%] | 16380<br>[80.8%] |
| Age | 75.6<br>(7.86) | 75.4<br>(7.79) | 75.3<br>(7.71) | 74.0<br>(6.91) | 73.9<br>(7.74) |
| Women | 1268<br>[51.8%] | 1270<br>[51.9%] | 1256<br>[51.7%] | 11398<br>[57.0%] | 11573<br>[57.1%] |
| Single households | 1011<br>[41.3%] | 1027<br>[42.0%] | 1072<br>[44.1%] | 9145<br>[45.7%] | 9372<br>[46.0%] |
| Finnish speaking | 557<br>[22.7%] | 526<br>[21.5%] | 544<br>[22.3%] | 17279<br>[86.4%] | 17423<br>[86.0%] |
| Swedish speaking | 1868<br>[76.2%] | 1898<br>[77.6%] | 1866<br>[76.8%] | 2087<br>[10.4%] | 2186<br>[10.8%] |

Note: This table summarizes descriptive baseline characteristics by region and treatment. The reported statistics are frequencies, except for variable *age* that shows the average age by region and treatment. Square brackets report proportions (%) and parentheses show standard deviation.

The average age in our samples was approximately 75 years. The majority of individuals were women and lived in households with two persons aged 65 and above. The Western Region is a bilingual region where the primary native language is Swedish (76·9%), whereas the Southern region is a largely monolingual region where the large majority of individuals (86·2%) are native Finnish speakers.

### Confirmatory analyses (pre-registered)

The primary analysis compares influenza vaccination coverage across the experimental arms in the Western and Southern regions. We report intention-to-treat results. Thus, individuals in all treatment arms were expected to remain in the initially assigned treatment group. The only potential sources of attrition were emigration or mortality after the postal address was extracted from the population register. There was no reason to expect that attrition would be correlated with treatment.

We first report the proportions and differences in proportions in influenza vaccination coverage by treatment arm in the Western and Southern regions (Figure 2). The statistical analysis adjusts for clustering at the household level. In the Western region, we observed the highest rate of vaccination coverage in the individual treatment (41·8%, 95% CI: 39·5% to 44·1%), the second highest rate of vaccination coverage in the individual + social treatment (38·9%, 95% CI: 36·6% to 41·2%), and the lowest rate of vaccination coverage in the no letter control condition (34·0%, 95% CI: 31·8% to 36·2%). The difference in proportions between the individual letter and no letter was 7·8 percentage points (95% CI: 4·6 pp to 11·0 pp, p < 0·001), between the individual + social letter and no letter 4·9 percentage points (95% CI: 1·7 pp to 8·1 pp, p = 0·002) and between the two letters 2·9 percentage points (95% CI: - 0·4 pp to 6·1 pp, p = 0·087). Finally, we pooled both information treatments (Figure 3A) and found that the effect of receiving any written information letter versus being in the control group without any written information was 6·4 percentage points (95% CI: 3·6 pp to 9·1 pp, p < 0·001).

**Figure 2:**
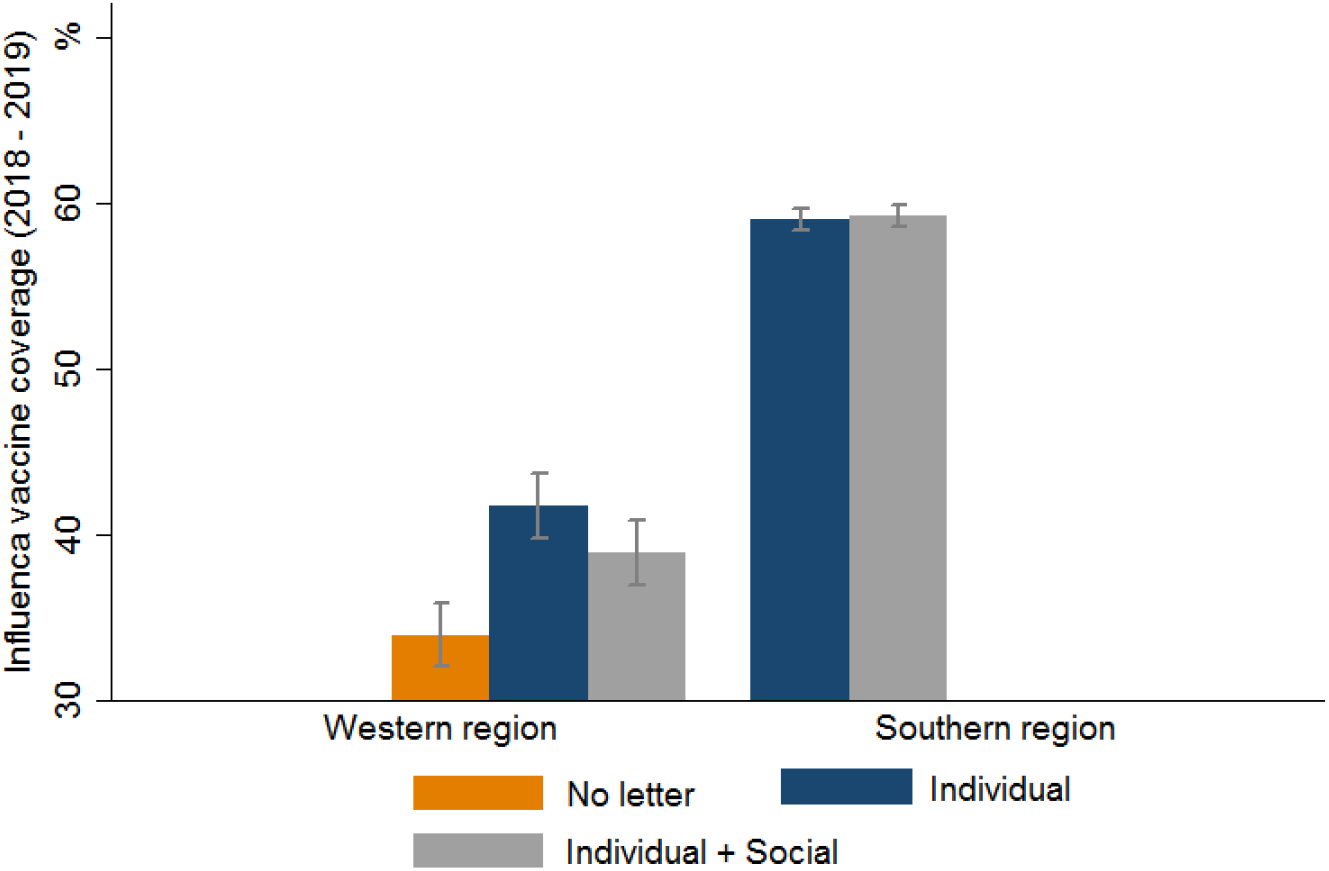
Vaccination coverage by treatment and region. Bar graphs denote influenza vaccination coverage. Error bars denote 95% confidence intervals

**Figure 3:**
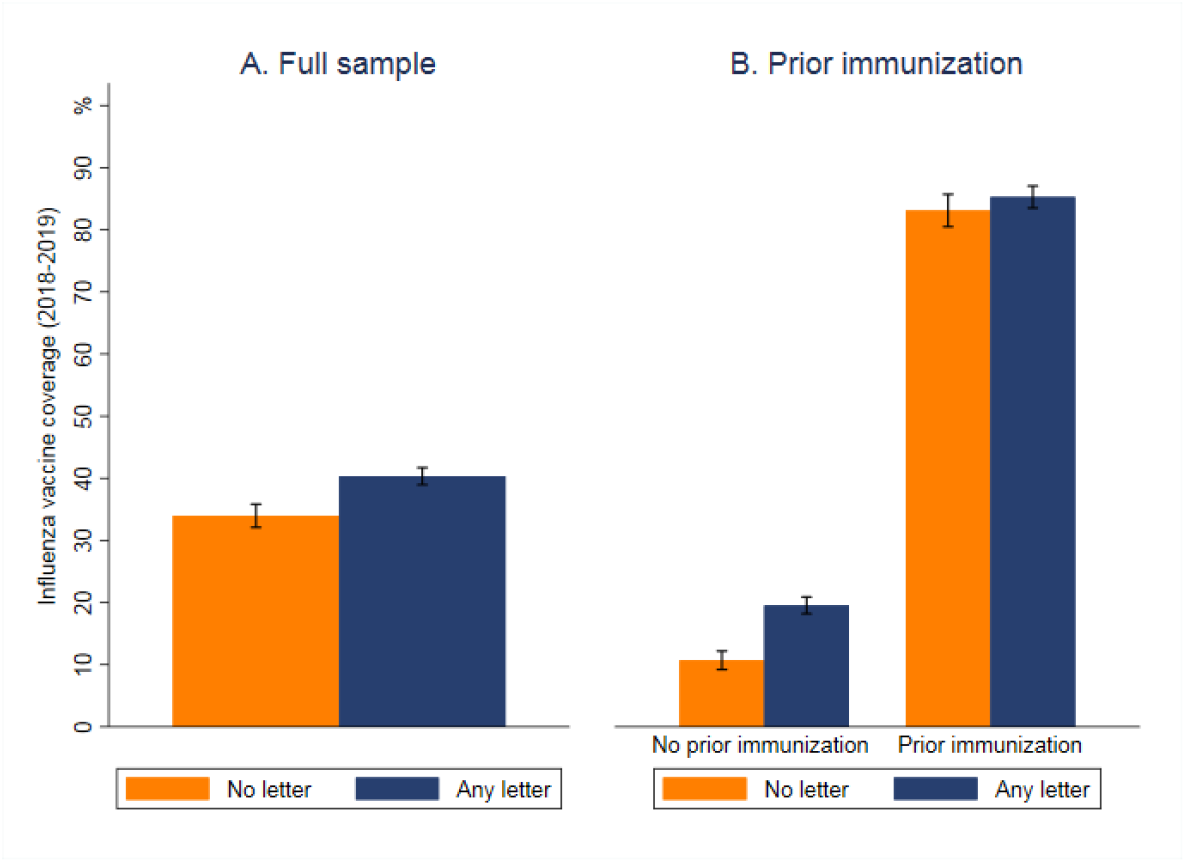
Vaccination coverage by treatment in the Western region. *Panel A:* Full sample (No letter vs. Any letter), *Panel B:* Vaccination coverage by treatment in the Western region stratified by prior vaccination status (No letter vs. Any letter). Error bars denote 95% confidence intervals.

In the Southern region, we observed that vaccination coverage was very similar in the individual and social benefit treatments (59·2%, 95% CI: 58·5% to 60·0%) and in the individual benefit treatment (59·0%, 95 CI: 58·3% to 59·8%). Consequently, the difference in proportions in vaccination coverage between the treatments I and I+S was very small (0·2 percentage points, 95% CI −0·09% to 1·30%, p = 0·724), indicating that there was no difference in vaccination coverage between the two treatments.

### Exploratory analyses (not pre-registered)

Our two administrative data sources enabled more exploratory analyses of heterogeneous treatment effects. We explored the effect of information letters conditional on prior vaccination history. Moreover, we estimated possible cross-vaccination spillovers from influenza vaccination to other common vaccinations among the age-group. Only data from the Western region entered the analyses as only this sub-design included a group of individuals who did not receive any letter.

A body of literature has documented that a variety of indirect suggestions and modified information disclosures (information nudges) can change behavior in a wide variety of contexts.^16,17^ However, there are an increasing number of findings documenting behavioral interventions that fail to influence behavior or change the behavior in the opposite direction than expected.^18,19^ Overall, little is known about the optimal design of information nudges in heterogeneous groups where individuals may have widely different beliefs about the potential risks and benefits of their actions or the desired behaviors. Here, we argue that prior vaccination history may serve as a proxy for positive or negative beliefs about vaccination and used administrative individual-level data on prior vaccination histories to investigate the effectiveness of information nudges conditional on prior vaccination history.

We estimated the treatment effect of our information intervention conditional on one of three indicators of the individual vaccination history: having vs. not having received influenza vaccination during the previous seasonal influenza period (2017 – 2018); having vs. not having received any influenza vaccination during the past nine years (from 2009 to 2018) prior to the influenza season 2018 – 2019; and having vs. not having received any vaccination during the past nine years (from 2009 to 2018) prior to the influenza season 2018 – 2019. The length of the prior vaccination period (nine years) was based on data availability and maximizes the available length of individual vaccination histories before the treatment assignment.

Table 2 (columns 1 – 2) shows the joint effect of any letter versus no letter, conditional on influenza vaccination status during influenza season 2017 – 2018 (one year prior to the study). We found that the effect of receiving any (vs. no) letter on vaccination coverage was substantially larger among the previously non-vaccinated individuals (8·8 percentage points higher in the letter vs. no letter conditions, which corresponded to a relative increase of 82 percent) than among the previously vaccinated individuals (2·2 pp increase).

**Table 2:** The effect of written information letters on influenza vaccination coverage by prior vaccination history

|  | Influenza vaccine uptake |  |  |  |  |  |
| --- | --- | --- | --- | --- | --- | --- |
|  | (1) | (2) | (3) | (4) | (5) | (6) |
| Any letter | 0.022<br>(.017) | 0.088***<br>(.017) | 0.048**<br>(.019) | 0.084***<br>(.011) | 0.050***<br>(.017) | 0.053***<br>(.013) |
| Coverage in control group (%) | 83.1% | 10.7% | 68.0% | 6.3% | 45.5% | 5.0% |
| Observations | 2329 | 4995 | 3243 | 4081 | 5308 | 2016 |
| Influenza vaccine, 2017 - 2018 | Yes | No | - | - | - | - |
| Prior influenza vaccine | - | - | Yes | No | - | - |
| Any prior vaccine | - | - | - | - | Yes | No |

Columns 3 and 4 shows the joint effect of any letter versus no letter, conditional on having vs. not having received influenza vaccination during the past nine years. We found that receiving vs. not receiving written letter increased vaccination coverage by 8·4 percentage points among individuals who had not received any influenza vaccination during the previous nine years. For those who had received at least one vaccination during the past nine years, the increase was 4·8 pp.

Columns 5 and 6 show the effects of receiving vs. not receiving a letter among those who have vs. have not received any type of vaccination during the previous nine years. We find a substantial positive effect (5·3 percentage points) even among the consistently non-vaccinated individuals. As the overall influenza vaccination coverage is low (5·0%) in this unvaccinated group, the relative effect size of receiving any letter is very large among the consistently non-vaccinated individuals (106 percent).

Finally, we examined whether receiving an individual letter reminding about the importance of influenza vaccinations increased vaccination coverage for other common vaccinations among the target population. These analyses utilized the fact that our data included comprehensive patient records about all vaccinations received after the implementation of the intervention. We estimated cross-vaccination spillovers separately for three most common types of vaccinations, other than the influenza vaccinations, in this age-group: pneumococcal conjugate vaccine (PCV), tetanus-diphtheria vaccine (TD) and tick-borne encephalitis vaccine (TBE). Moreover, we estimated the effect of receiving a written information letter on the receipt of any other vaccine than influenza vaccine. Our results are reported in the online appendix D and strongly indicate that there were no cross-vaccination spillovers. The estimated effects in all models were bounded to a tight interval around zero.

## Discussion

Public health authorities across the world recommend yearly influenza vaccinations to individuals who are at high risk for severe disease. However, the uptake of influenza vaccinations remains low, despite the recommendations, educational efforts, financial subsidies and reliable supply of influenza vaccination services. The aim of this paper was to understand the behavioral determinants of individual vaccination decisions and investigate the effectiveness of behavioral interventions that may complement more conventional approaches to improve the uptake of influenza vaccination.

This paper presents new causal evidence on the factors that affect influenza vaccination decisions among elderly population. Our paper extends the study of behavioral interventions from hypothetical vaccination intentions and small-scale outpatient settings to a large-scale cluster-randomized controlled trial in which vaccination decisions are measured using comprehensive health records that include information about all received vaccinations before and during the follow-up period. More generally, this study serves as an example how a randomized study design can be merged to high-quality administrative data to estimate causal effects in large and representative samples.

We observed that a low-cost and scalable intervention relying on individually mailed letters substantially increases influenza vaccination coverage. Comprehensive patient records enabled us to condition the effect of mailed letters on individuals’ prior vaccination history. The analyses revealed that the effect of information letters was substantially larger among the individuals who had not received influenza vaccination in the previous year. Moreover, we observed that even the most consistently non-vaccinated individuals, who had not received any vaccination during the previous nine years, had a positive response to a written reminder. In contrast, there was no statistically significant effect among previously vaccinated individuals. These findings suggest that the cost-effectiveness of interventions aiming to improve vaccination coverage will crucially depend on the prior vaccination history of the target population. Overall, using comprehensive and exact administrative information about prior vaccination histories, or statistical variables that predict prior vaccination history in the absence of exact health care records, can provide a promising avenue to enhance the effectiveness of behavioral interventions aiming to improve vaccination coverage.

Our results suggest that a written explanation about the social benefits of vaccinations in addition to the individual benefits did not increase influenza vaccination coverage. More generally, we found that appealing to collective responsibility in addition to the decreasing complacency does not affect influenza vaccination coverage. Consequently, we conclude that, at least in the case of influenza vaccination and for the letter intervention used, communicating the social benefits of vaccination due to herd effect leads neither to pro-social vaccination nor free-riding on the vaccination efforts of other community members.

The present results are largely consistent with the literature that has documented the effectiveness of patient reminders and recall interventions on vaccination coverage.^20-23^ However, the vast majority of existing evidence stems from outpatient provider office settings in which there are an active care relationship between the provider and patient. The conclusions from these studies may not necessarily apply to large-scale interventions within the general elderly population. In contrast, our study overcomes these limitations and tests the effectiveness of information letters as an easily scalable and low-cost communication strategy in a general elderly population.

This paper is related to a nascent literature that has tested the effectiveness of various communication strategies and behavioral interventions on vaccination coverage across different vaccinations and populations.^24-26^ There is also recent evidence suggesting that communicating the social-benefit motives of vaccinations to hospitalized high-risk patients does not enhance actual vaccination behavior.^27^ Our null results related to the effects of communicating the social benefits of vaccination parallel the emerging conclusion from the literature that information materials tailored using behavioral science techniques do not affect real vaccination rates. In contrast, there is some evidence from the low- and high-income countries that modest in-kind incentives and direct monetary incentives may increase vaccine uptake.^28,29^ Overall, it still remains to be studied whether communicating the social benefits of vaccination due to herd effect increases vaccine uptake against more contagious diseases, such as measles, which have a clear threshold for herd immunity.

We acknowledge that our study has several limitations. First, there could have been some contamination between the treatments if information about the letters and their content was shared between individuals (e.g. neighbors, friends, and other individuals in receiver’s social networks) who belonged to different treatment groups. However, these kinds of information spillovers were minimized by the cluster-randomized design that guaranteed same information for the household members. Second, the effectiveness of information letters may be underestimated. This study reports intention-to-treat effects that disregard questions about the effectiveness of letters among individuals who actually opened and read the letters. While the generally very efficient and reliable postal services in Finland increase confidence that the letters were delivered to the households in due time, we do not have information about the proportion of letters that were opened and read by the individuals. Here, the fact that the letters were written on a paper that included a printed letterhead and were signed by the local chief physicians have likely minimized recipients’ potential concerns about the authenticity of letters. Third, we are not able to identify the impact of information letters on influenza vaccination coverage per se in the Southern region because all individuals in the study population were assigned either to the individual benefit treatment or to the individual and social benefit treatment. Thus, we are not able to infer whether the effect of receiving any letter depends on the aggregate rate of vaccination coverage in the study population. Fourth, the effects of communicating the social benefits of vaccination due to herd effect may depend on the description and presentation of social benefits. This study is limited to describing the mechanism of herd effect in a written format using a layout that could be easily fit into a standard information letter. It remains to be tested whether communicating the social benefits of vaccination using other communication formats (e.g. graphical presentation or moving images) would affect real vaccination decisions.

In conclusion, this large-scale cluster-randomized controlled trial has shown how a behavioral intervention study can be combined with routinely collected high-quality administrative data to estimate causal effects in large and representative samples. We observed that a letter reminder informing elderly individuals about the individual benefits of vaccination led to a substantial increase in influenza vaccination coverage. This positive effect on influenza vaccination coverage was observed even among the most consistently non-vaccinated individuals. Informing individuals about the social benefits of vaccination did not further increase vaccination coverage. These findings have meaningful implications for the financing of preventive health interventions and public health authorities who implement vaccination communication strategies to enhance vaccine uptake and aim to curb the spread of infectious diseases.

## Data Availability

De-identified individual-level data will be shared using the Open Science Framework data repository. Data sharing will commence immediately following publication. Statistical code to organize the data and replicate the statistical analysis is made available using the Open Science Framework data repository.

## Contributors

LS wrote the manuscript, participated in the design of the study, collected data and performed the statistical analyses. CB and RB participated in the design of the study, the data analysis plan and revising of the manuscript. JS, HS and HN participated in design of the study, collection of data and revising of the manuscript. All authors agreed on the content of the manuscript, reviewed drafts, and approved the final version

## Supplementary Material

### 1. English Translation

#### Invitation to Influenza Vaccination!

We would like to invite all citizens aged 65 and over living in Coastal Ostrobothnia (Maalahti, Korsnäs, Närpiö, Kaskinen and Kristiinankaupunki) for free influenza immunization.

Seasonal influenza is a common and serious disease in the age group of 65 and above. Influenza vaccination is the best way to protect you against the disease. Influenza vaccine will protect you also against many secondary diseases associated with seasonal influenza such as pneumonia. It is recommended to take an influenza vaccine every autumn as the protective effect of these vaccines last for about a year. Influenza viruses continuously change and previously taken vaccines may not provide protection in the following years.

**You may receive your influenza vaccine without appointment at following dates and times Kristiinankaupunki - Children’s health clinic (Address: Lapväärtintie 10)**

- **Monday 29.10. from 1 p.m. to 5 p.m**.
- **Monday 05.11. from 1 p.m. to 5 p.m**.
- **Monday 26.11. from 1 p.m. to 5 p.m**.
- **Monday 10.12. from 1 p.m. to 5 p.m**.

**Siipyy - Children’s health clinic (Address: Långvikintie 16)**

- **Wednesday 31.10. from noon to 3 p.m**.
- **Wednesday 14.11. from noon to 3 p.m**.

**You may also book an appointment for the vaccine administration on weekdays by calling 06 221 8480**

Please bring you social security card with you. We recommend wearing clothes that enable injection of *a vaccine into the shoulder*.

**Please notice that if you receive medical home care or live in a nursing home**, you may receive an influenza vaccine directly through your care givers. You do not have to book an appointment and travel to receive your vaccine.

**Welcome**

**Peter Riddar**, Chief physician

For additional information please contact: www.kausi-influenssa.fi

Your mail address was extracted from the Population Register, Population Register Center, P.0. Box 123, 00531 Helsinki This invitation has been prepared in cooperation with the Finnish Institute for Health and Welfare.

### 2. Randomization Script (Appendix B)

*Data management after receiving the address data

import excel “\\helfs01.thl.fi\cData\Rokoteviesti_RCT\Tulokset 2018-09-28.xlsx”, sheet(”Tulostiedot”) firstrow

********************************************************************************

*KEEP only Kaskinen

keep if Kunnannimi==”Kaskinen”

*Create unique identifiers by apartment

sort Asuinpaikantunnus

egen running_apartment = group(Asuinpaikantunnus)

******************************************************************************

bys Asuinpaikantunnus: gen n_of_persons = _N

******************************************************************************

set seed 21042403

gen random_number =uniform()

bysort Asuinpaikantunnus: replace random_number = random_number[1]

egen ordering = rank(random_number), unique

gen treatment = “”

*Control treatment

replace treatment = “C” if ordering <= _N/3

*Standard treatment

replace treatment = “TS” if ordering <= 2*_N/3 & ordering > _N/3

*Herd treatment

replace treatment = “TH” if ordering <= 3*_N/3 & ordering > 2*_N/3

### 3. Power calculations (Appendix C)

The implementation of our randomized controlled trial was guided by an aim to run the study among the entire elderly population (≥ 65 years of age) in two different samples. The randomization took place at the household (cluster) level to avoid sending letters with different contents to same household members. These two practical design principles determine our total sample size and the number of clusters (households) in our samples. Using information on sample sizes and number of clusters, we can report the minimum detectable effect size (MDE) for different treatment effects and assess whether our null findings identify the absence of a true effect or signify a lack of statistical power.

The MDE is a metric to measure the smallest effect that would have been detectable given our samples sizes and clusters. Here we compute the MDEs with α = 0.05 and 0.80 power using different intracluster correlations for each pairwise comparison of our treatments. We note that the practice of reporting MDEs is substantially more conservative than simply stating the bounds of the 95% confidence interval.

**Figure A1:**
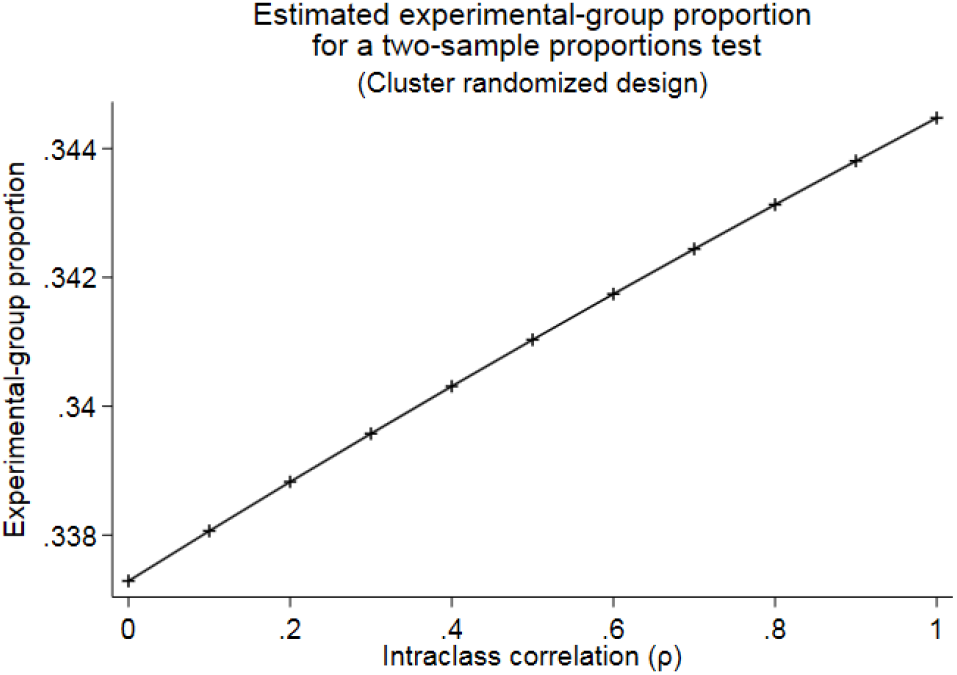
Minimal detectable effect sizes (with α = 0.05 and 0.80) for treatment comparisons by intracluster correlation coefficients in the Western region

#### Western region – treatment comparisons

Our analysis sample includes 7324 (5221 clusters) individuals that are randomized into three different treatments: Control treatment (N = 2450, 1732 clusters), Individual Treatment (N = 2445, 1738 clusters) and Individual and Social Treatment (N = 2429, 1751 clusters). Using a baseline immunization rate of 30% and assuming 80% power for a 5% (two-sided) level test, we determine that a sample size of 2450 individuals and 1732 clusters yields a minimal detectable effect size that varies from 3.7 percentage points to 4.5 percentage points depending on the intraclass correlation within clusters (Figure A1).

**Figure A2:**
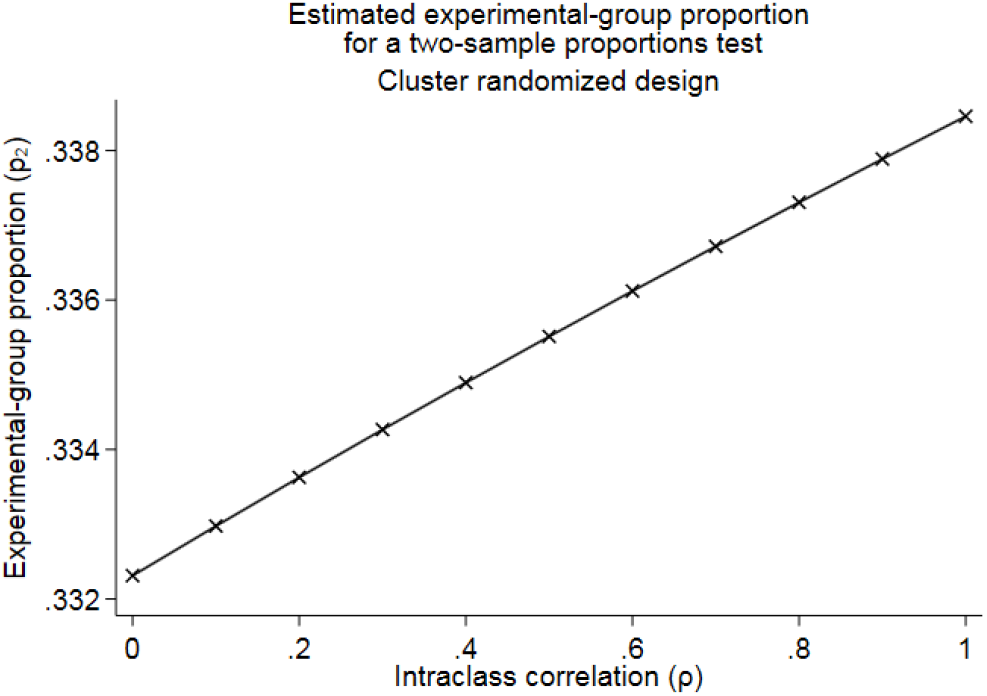
Minimal detectable effect sizes (with α = 0.05 and 0.80) for the joint effect of any letter s by intracluster correlation coefficients in the Western region

#### Western region – The effect of any letter

Our analysis sample for estimating the impact of an information letter *per se* includes 7324 (5221 clusters) individuals that are divided into Treatment - any letter (N =4874, 3489 clusters) and control treatment (N = 2450, 1732 clusters). Using a baseline immunization rate of 30% and assuming 80% power for a 5% (two-sided) level test, we determine that this treatment comparison yields a minimal detectable effect size that varies from 3.2 percentage points to 3.8 percentage points depending on the intraclass correlation within clusters. (Figure A2).

#### Southern region – treatment comparison

Our analysis sample includes 40271 (29395 clusters) individuals that are randomized into two different treatments: Treatment A (N = 19996, 14571 clusters) and Treatment B (N = 20275, 14824 clusters). Using a baseline immunization rate of 58% and assuming 80% power for a 5% (two-sided) level test, we determine that this treatment comparison yields a minimal detectable effect size that varies from 1.4 percentage points to 1.6 percentage points depending on the intraclass correlation within clusters (Figure A3).

**Figure A3:**
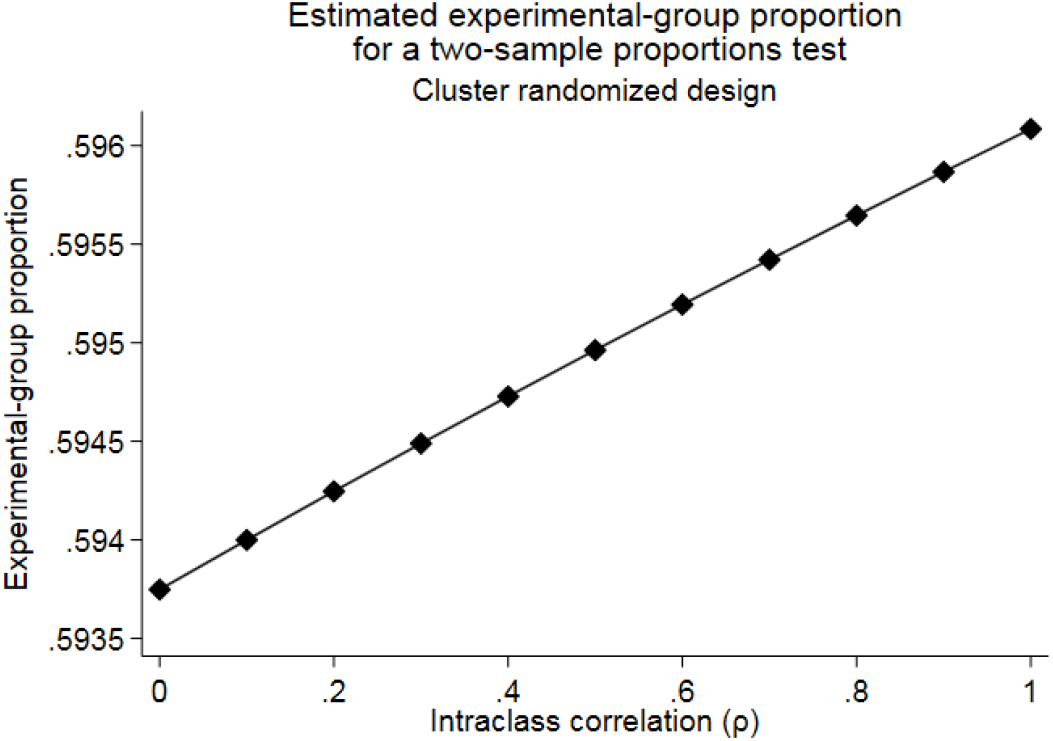
Minimal detectable effect sizes (with α = 0.05 and 0.80) for the treatment comparison by intracluster correlation coefficients in the Southern region

### 4. Supplementary Figures and Tables (Appendix D)

#### 4.1 Robustness

**Table A1:** Average treatment effects estimated using linear probability models, logit models and generalized mixed effects regression with random effects

| Western Region |  | Southern Region |  |
| --- | --- | --- | --- |
| Diff. Any letter<br>– Control group | Diff. Treatment I<br>– Treatment I+S | Diff. Any letter<br>– Control group | Diff. Treatment I<br>– Treatment I+S |
| Linear probability model |  |  |  |
| 6.4 pp | 2.9 pp | - | - 0.2 pp |
| [3.6 pp – 9.1 pp] | [0.0 pp – 6.1 pp] | - | [-1.3 pp – 0.9 pp] |
| (P < 0.000) | (P = 0.087) | - | (P = 0.724) |
| Logit – Marginal effects |  |  |  |
| 6.4 pp | 2.9 pp | - | - 0.2 pp |
| [3.6 pp – 9.1 pp] | [0.0 pp – 6.1 pp] | - | [-1.3 pp – 0.9 pp] |
| (P < 0.000) | (P = 0.087) | - | (P = 0.724) |
| Random effects model |  |  |  |
| 6.3 pp | 2.6 pp | - | - 0.1 pp |
| [3.7 pp – 9.0 pp] | [-0.1 pp – 5.7 pp] | - | [-1.2 pp – 1.0 pp] |
| (P = 0.000) | (P = 0.107) | - | (P = 0.840) |

Notes: This table reports means by treatment and differences between treatments. Squared brackets show 95% confidence intervals. P-values refer to the test of difference between treatments. To take into account the clustered randomization design linear probability models and logit models use standard errors clustered at household level, random effect model includes household as a random intercept.

**Table A2:** The effect of written information letters on influenza vaccine take-up by prior immunization history – Random effects linear model

|  | Influenza vaccine take-up |  |  |  |  |  |
| --- | --- | --- | --- | --- | --- | --- |
|  | (1) | (2) | (3) | (4) | (5) | (6) |
| Any letter | 0.024<br>(.018) | 0.089***<br>(.112) | 0.052***<br>(.019) | 0.088***<br>(.012) | 0.060***<br>(.016) | 0.053***<br>(.014) |
| Sample mean (%) | 84.5 | 16.7 | 71.3 | 11.9 | 49.5 | 8.5 |
| Observations | 2329 | 4995 | 3243 | 4081 | 5308 | 2016 |
| Influenza vaccine 2018 | Yes | No | - | - | - | - |
| Prior influenza vaccine | - | - | Yes | No | - | - |
| Any prior vaccine | - | - | - | - | Yes | No |

Notes: Reported coefficients are derived using random effects linear models that allow cluster level heterogeneity (random effects) at the household level. All regressions are estimated at the individual level. Parentheses present standard errors from the random effects models. \*\*\*Significant at $p < 0.01$ , \*\*Significant at $p < 0.05$ , \*Significant at $p < 0.1$ .

**Table A3:** The effect of written information letters on influenza vaccine take-up by prior immunization history – Logit model (marginal effects)

|  | Influenza vaccine take-up |  |  |  |  |  |
| --- | --- | --- | --- | --- | --- | --- |
|  | (1) | (2) | (3) | (4) | (5) | (6) |
| Any letter | 0.022<br>(.017) | 0.089***<br>(.012) | 0.048**<br>(.019) | 0.084***<br>(.011) | 0.060***<br>(.017) | 0.053***<br>(.013) |
| Sample mean (%) | 84.5 | 16.7 | 71.3 | 11.9 | 49.5 | 8.5 |
| Observations | 2329 | 4995 | 3243 | 4081 | 5308 | 2016 |
| Influenza vaccine 2018 | Yes | No | - | - | - | - |
| Prior influenza vaccine | - | - | Yes | No | - | - |
| Any prior vaccine | - | - | - | - | Yes | No |

Notes: Reported coefficients are derived using logit models (marginal effects) random effects linear model. All regressions are estimated at the individual level. Parentheses present standard errors clustered at the household level. \*\*\*Significant at $p < 0.01$ , \*\*Significant at $p < 0.05$ , \*Significant at $p < 0.1$ .

#### 4.2 Cross-vaccine spillovers

We examined whether receiving a letter on influenza vaccination increases vaccine uptake for other common vaccines among our target population group. These types of cross-vaccination spillovers may occur through several behavioral channels. First, the letters could increase the interaction between citizens and health care personnel who administer vaccinations. This interaction may lead to information exchange where providers inform citizens about the possibility of receiving other age-appropriate vaccinations. Second, the letters may encourage individuals to gather knowledge about other available age-appropriate vaccinations from other available information sources (e.g. internet, books, and brochures). Third, the letters may lead to changes in perceived confidence in vaccination in general and alter the inclination to take available age-appropriate vaccines.

We utilize the fact that our data included comprehensive patient records about all vaccines received after the implementation of the experiment. We estimate cross-vaccination spillovers in the Western region separately for pneumococcal conjugate vaccine (PCV), diphtheria vaccine (DV) and tick-borne encephalitis vaccine (TBE).

**Table A4:** Cross-vaccination spillovers to other age-appropriate vaccines

|  | PCV | DT | TBE | Any vaccine |
| --- | --- | --- | --- | --- |
|  | (1) | (2) | (3) | (4) |
| Any letter | 0.001<br>(.004) | 0.001<br>(.003) | 0.001<br>(.001) | -0.001<br>(.005) |
| Sample mean (%) | 0.5% | 1.6% | 0.2% | 3.9% |
| Observations | 7,324 | 7,324 | 7,324 | 7,324 |
| Notes: Reported coefficients are derived using linear probability models. All regressions are estimated at the individual level. Standard errors in parentheses are clustered at the household level. ***Significant at $p < 0.01$, **Significant at $p < 0.05$, *Significant at $p < 0.1$. | | | | |

Table A4 presents the cross-vaccination spillovers. The first column shows the effect on PCV, second column on DT, third column on TBE vaccination. The fourth column shows the effect of having received any of the three vaccinations. There were no cross-vaccination spillovers. Using the 95% confidence intervals, we are able to rule out for PCV, DT, and TBE effects smaller than 0.5 percentage points and larger than 0.7 percentage points. For any vaccine, we can rule out effects smaller than −1.1 percentage points and larger than 0.9 percentage points. Overall, we find that information letters do not cause any cross-vaccination spillovers.

